# The effectiveness of healthy lifestyle training in migrant domestic helpers in Hong Kong: a quasi-experimental study

**DOI:** 10.64898/2025.12.15.25342332

**Authors:** Jie V Zhao, Bing Chen, Michael Magtoto Manio

**Affiliations:** School of Public Health, Li Ka Shing Faculty of Medicine, the University of Hong Kong; School of Biomedical Sciences, Li Ka Shing Faculty of Medicine, the University of Hong Kong

**Author notes:** Correspondence, 2/F, Patrick Manson Building, 7 Sassoon Road.

## Abstract

**Background:** Migrant domestic helpers are vulnerable to obesity and mental disorders, yet few interventions target this population. To address the research gap, we designed healthy lifestyle intervention tailored to this population and evaluated its effectiveness on healthy behaviors and mental well-being among Filipino domestic helpers in Hong Kong.

**Methods:** The intervention included dietary education and yoga practice, delivered onsite or online. Questionnaire surveys were conducted at baseline, immediately post-intervention, and four weeks post-intervention. Healthy Eating Vital Sign (HEVS) assessed dietary behvior, and PHQ-4 assessed mental health. Linear mixed-effects models were used to assess the changes in dietary behaviors and mental health.

**Results:** Of 318 participants (271 onsite), over 95% reported willingness to improve diet and lifestyle, and recommend the program to employer families. Four weeks post-intervention, participants showed statistically significant improvements in dietary behavior (HEVS score: - 0.55; 95% confidence interval (CI): -0.92 to -0.18) and mental health (PHQ-4 score: -0.30; 95% CI: -0.57 to -0.02), with greater benefits among those with poorer baseline diet (HEVS: -1.90; 95% CI: -2.50 to -1.29) or higher mental stress (PHQ-4: -0.71; 95% CI: -1.16 to -0.27). Improvements included vegetable and fruit intake, breakfast frequency, and reduced depressive symptoms.

**Conclusions:** This healthy lifestyle intervention was well-accepted and brought improvements in dietary behavior and mental health among migrant domestic helpers, particularly those at higher risk. These findings contribute to an under-researched area and highlight the potential of integrating such low-cost, accessible interventions into community health initiatives to reduce health disparities in this vulnerable population.

## Introduction

The health and well-being of transnational migrant domestic helpers is an urgent yet largely overlooked public health issue.^1^ The Asia-Pacific region accounts for more than 20% of the world’s migrant domestic helper population.^1^ Hong Kong is a major destination,^2^ driven by its relatively high wages and strong legal protections.^3^ By 2024, the number of domestic helpers in Hong Kong reached approximately 368,000, representing nearly 10% of the city’s workforce.^2^ Since the introduction of the foreign domestic helper policy in the 1970s,^4^ these workers have become an essential part of Hong Kong society. Beyond housekeeping, they take on roles as cooks, child caregivers, elderly care providers, and household managers, enabling dual-income families to remain in the workforce.^2^

Despite their essential role, domestic helpers remain a vulnerable group. Many work 12-16 hours a day with blurred boundaries between work and rest, at the live-in arrangements.^5, 6^ Poor dietary habits are common, with nearly 64% of Filipino domestic helpers in Macau classified as overweight or obese, more than double the 30.8% rate among adults in the Philippines.^7^ Separation from family, migration status, structural and cultural barriers, and lack of social support make them vulnerable to emotional stress,^3, 5, 8-10^ with depression affecting up to 25% of Filipino domestic helpers in Hong Kong^11^ and 40% in Macau.^12^ The challenge is exacerbated by structural and practical barriers restricting their access to and utilization of healthcare resources.^3, 8, 13, 14^ All these factors make them a critical population for public health intervention.

Healthy diet and yoga practice are well-established interventions known to improve cardiometabolic health and reduce stress.^15^ However, we found no studies evaluating their effectiveness among migrant domestic helpers. In fact, very few studies have examined health promotion specifically for this population. One quasi-experimental study involving 88 Filipino domestic helpers in Macau showed that online health literacy education was feasible and improved participants’ health literacy.^16^ However, this study did not assess behavioral changes. Another study, a clinical trial in Singapore with 40 Filipino domestic helpers, evaluated paraprofessional training based on cognitive behavioral therapy to help manage depression.^17^ This trial reported no significant differences in primary outcomes, partly due to its small sample size.^17^ Given this research gap, we developed a tailored intervention combining healthy diet education and yoga practice for domestic helpers and examined its effectiveness in improving health behaviors and mental well-being.

## Methods

### Study design

In this study, we evaluated the effectiveness of a healthy lifestyle intervention that combines diet education and yoga practice. Data were collected at three time points: baseline (pre-intervention), immediately post-intervention, and four weeks post-intervenion. We also collected their feedback throughout the program. The primary objective is to examine the changes in dietary behaviors, and mental well-being, the secondary objective is to examine the changes in physical activity, the acceptability of the project and their willingness to share with the families they serve, among domestic helpers in Hong Kong.

### Study population and recruitment

This study focused on Filipino female domestic helpers, who constitute the majority (55%) of foreign domestic helpers in Hong Kong. Filipino and Indonesian helpers, while both essential to the community, have distinct cultural backgrounds. Considering this is the first study of this type of intervention, we conducted within this single, homogeneous group, before extending to other populations, such as Indonesians, in further studies. Specifically, the inclusion criteria are: (a) female domestic helpers, (b) currently working in Hong Kong, and (c) being able to understand and read English.

We recruited participants via poster delivery and promotion among local domestic helper employment agencies, and non-governmental organization (NGO) supporting domestic helpers, HELP for Domestic Workers. Eligible participants received a supermarket voucher (50 HKD) on study completion as an appreciation for their time. All participants were informed that participation is voluntary and that they can withdraw at any time without penalty.

### Study process and intervention

Participants who agreed to take part in the study were screened for eligibility based on the criteria described above. Once eligibility was confirmed, participants were invited to provide informed consent. Those who consented received a copy of the consent form along with contact details of the study team, ensuring they could reach staff at any time.

The intervention consisted of two components: healthy diet education and physical activity (yoga) training. All sessions were scheduled on Sundays, which is the participants’ regular day off. Training was delivered in two formats: onsite and online. The online training was hosted on a dedicated project website containing all training materials, designed to accommodate participants who were unable to attend onsite sessions.

For onsite training, each participant first completed a 10-15-minute session on healthy diet education, followed by a 30-minute supervised yoga practice. The healthy diet education covered fundamental concepts of health, principles of healthy eating, and practical cooking methods (e.g., reducing oil, sugar, and salt). It also included actionable tips for applying these principles in daily life.

The yoga component was tailored specifically for domestic helpers, featuring standing postures that could be practiced during short breaks between tasks. Participants first watched a video demonstration and then practiced under the guidance of a qualified yoga instructor. They were also provided access to the project website for self-review and continued practice.

For participants attending onsite training, questionnaire surveys were conducted at three time points: baseline (pre-intervention), immediately after the intervention, and four weeks post-intervention. In the baseline and 4-week post-intervention surveys, we assessed dietary behaviors, depressive symptoms and physical activity. In the second questionnaire administered immediately after the intervention, participants were asked about their willingness to change dietary behavior, whether they liked the yoga program, and whether they were willing to recommend the training to the families they serve.

For participants attending online training, only the second questionnaire was conducted, as we were unable to monitor the timing of their baseline responses and the timing when they finish the training. Feedback from all participants was collected throughout the program to inform feasibility and acceptability.

### Outcome assessment

Dietary behaviors were assessed using the Healthy Eating Vital Sign (HEVS),^18^ a validated 12-item questionnaire designed to evaluate dietary behaviors associated with overweight and obesity. HEVS includes items on behaviors such as fast-food consumption and intake of sugar-sweetened beverages. Compared to comprehensive dietary assessment tools like the Food Frequency Questionnaire, HEVS is shorter and easier to administer, making it well-suited for community-based interventions and large-scale studies.

Mental wellbeing was assessed using the Patient Health Questionnaire-4 (PHQ-4), a screening tool that combines two items for depression (PHQ-2) and two items for anxiety (GAD-2).^19^ PHQ-4 provides a rapid assessment of psychological distress and is widely used in both research and clinical settings.

Physical activity was measured using the International Physical Activity Questionnaire (IPAQ), a widely recognized instrument for assessing activity levels.^20^ IPAQ collects information on the frequency and duration of physical activities. It also distinguishes between different intensities of physical activity, such as walking, moderate activity, and vigorous activity.

### Statistical analysis

For dietary behavior, HEVS scores were coded such that higher scores indicated less healthy dietary behaviors, with possible values ranging from 1 to 7 for each item. The total score (range: 7-49) was derived by summing responses reflecting participants’ typical dietary behaviors over one day or one week, as reported in the questionnaire, and did not include single-day intake such as yesterday’s. For mental health, each item of PHQ-4 score ranged from 0 to 3, the total score ranged from 0 to 12, where higher scores reflected worse mental health status. Physical activity was analyzed based on self-reported days, hours, or minutes spent in physical activity during the previous week.

We used a linear mixed effects model to assess changes between baseline and 4 weeks post-intervention for each outcome (HEVS score, mental health and physical activity). The model included time (baseline vs. 4 weeks) as a fixed effect, along with age and education level as covariates. A random intercept for participant ID was included to account for repeated measures within individuals. Results are presented as estimated mean differences with 95% confidence intervals.

Considering that participants with different baseline levels of health behaviors and mental stress may respond differently to the intervention, we also conducted stratified analyses. Participants were divided into two groups for each variable based on the median of their baseline HEVS score, PHQ-4 score, or time spent on physical activity.

To assess program acceptability and participants’ willingness to change, we summarized the percentage of participants reporting willingness to change dietary behavior, the percentage who liked the yoga program, and the percentage of participants willing to recommend the training to the families they serve.

All the analyses were conducted using R (version 4.3.2, R Foundation for Statistical Computing, Vienna, Austria).

### Ethical approval

This study was approved by Institution Review Board of The University of Hong Kong / Hospital Authority Hong Kong West Cluster (approval number UW 24-823).

## Results

A total of 723 female domestic helpers registered for the program. Among them, 318 completed the training, with 85.2% attending onsite sessions and 14.8% participating online. The mean age of the 318 participants was 43.0 years, and 36.2% had completed high school education, 46.6% held undergraduate degree or higher. For those who completed onsite training (n□=□271), the median HEVS total score at baseline was 18, and the median PHQ-4 score was 1.

Immediately after the intervention, 97.4% of participants indicated willingness to change their diet and daily lifestyle, 95.9% expressed willingness to recommend our healthy diet education to their employer families, and 86.5% would like to recommend our yoga program to their employer family. Furthermore, 99.3% reported satisfaction with the yoga program. Among those who completed online training (n = 47), similar results were observed, with 97.8%, 97.9%, 93.6%, and 97.9%, respectively, for the above indicators.

Using linear mixed effects model, the participants showed a statistically significant improvement in dietary behavior. The HEVS total score decreased by an average of 0.55 units four weeks post-intervention compared to baseline (95% CI: -0.92 to -0.18). Mental health also improved, with the PHQ-4 total score decreasing by an average of 0.30 points (95% CI: -0.57 to -0.02). Notably, the benefits were more pronounced among participants with poorer baseline dietary behavior (estimate for HEVS: -1.90, 95% CI: -2.50 to -1.29) or higher mental stress (estimate for PHQ-4: -0.71, 95% CI: -1.16 to -0.27) (Figure 1).

**Figure 1.**
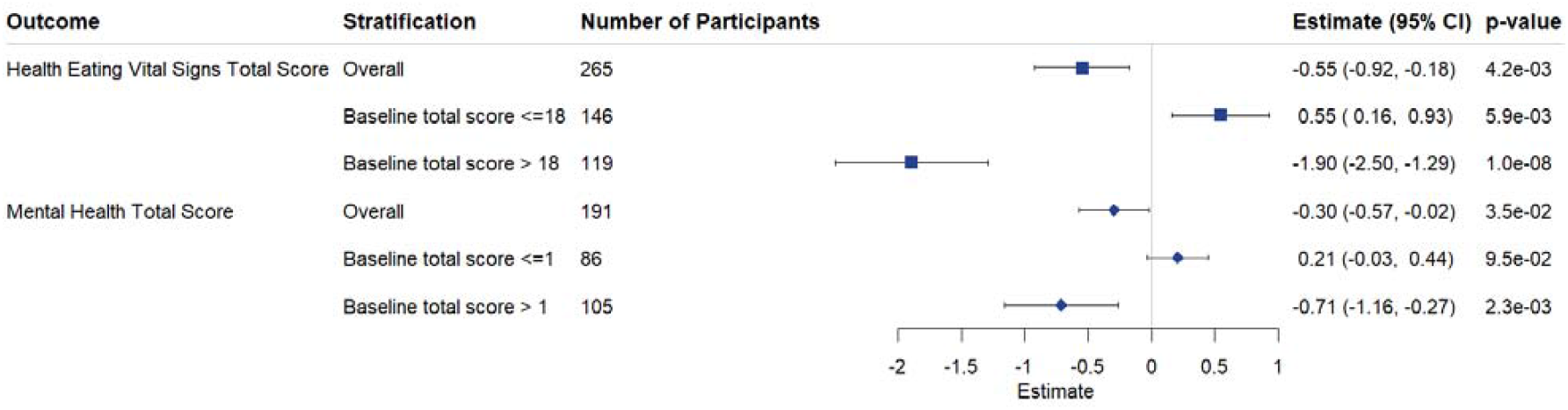
The changes in HEVS and PHQ-4 total scores at 4-week post-intervention, using linear mixed effects model

Further analysis of specific HEVS items among participants with greater improvements showed notable changes in vegetable intake, fruit consumption, and frequency of eating breakfast (Figure 2). For PHQ-4 items, improvements were observed in interest or pleasure in doing things and feeling down, depressed, or hopeless (Figure 3). Other mental health items showed downward trends after training, although these changes did not reach statistical significance (Figure 3).

**Figure 2.**
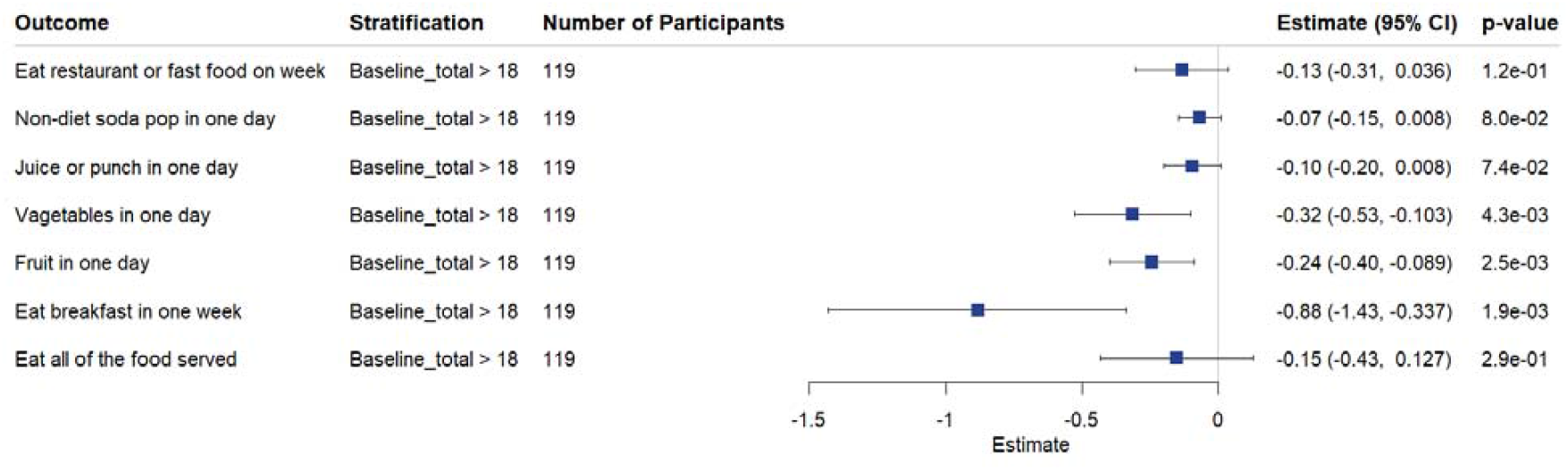
The changes in specific HEVS item at 4-week post-intervention, using linear mixed effects model among domestic helpers with poorer baseline dietary behavior

**Figure 3.**
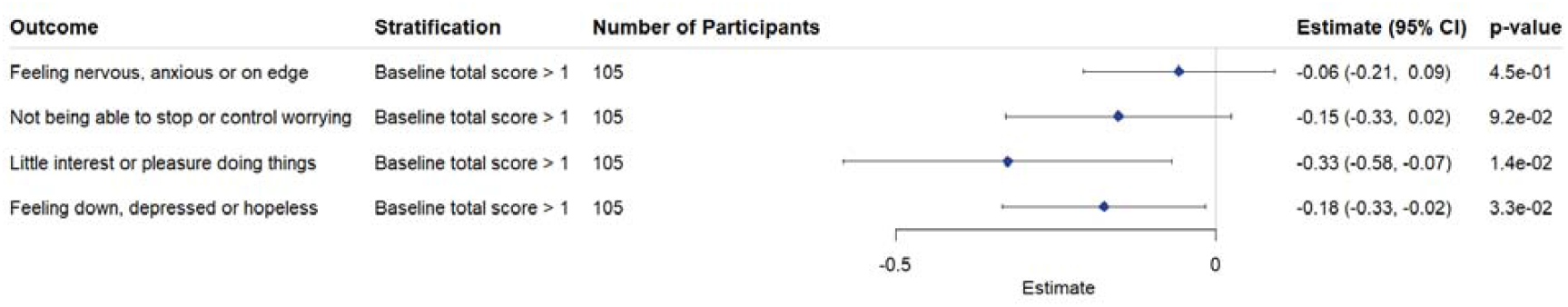
The changes in specific PHQ-4 item at 4-week post-intervention, using linear mixed effects model among domestic helpers with higher mental stress

For physical activity, no significant changes were observed in vigorous activity, moderate activity, walking, or sitting time in overall people. However, among participants with lower baseline physical activity, improvements were evident (Supplemental Figure 1). These participants reported more days per week engaging in vigorous activity, moderate activity, and walking for at least 10 minutes at a time during the follow-up period.

Feedback from the participants showed strong appreciation for the program, describing it as helpful, practical, and easy to follow. Several reported adopting healthier dietary behaviors, reducing intake of salty and processed foods, and incorporating yoga or stretching into daily routines. Several noted feeling more relaxed and motivated to prioritize their health and shared intentions to recommend the program to others. These comments highlighted high satisfaction and perceived benefits for both physical and mental well-being.

## Discussion

In the situation where few studies have evaluated health promotion interventions among migrant domestic helpers, our findings make an important contribution to this under-researched area. Despite the inconsistent evidence regarding the effectiveness of healthy diet intervention on dietary behaviors in general population,^21, 22^ our results suggest that in this marginalized group, healthy diet education can lead to measurable improvements in dietary behaviors, which adds to the limited evidence in the underrepresented group.^21^ Our findings also showed that yoga-based practices may reduce psychological distress. The observed improvements in HEVS and PHQ-4 scores align with literature demonstrating the benefits of lifestyle interventions on mental health outcomes in other populations.^15, 23^ The mental health improvements, particularly the increase in pleasure in doing things, align with qualitative feedback indicating that participants enjoyed the yoga sessions and found them relaxing and uplifting.

Our findings showed high feasibility and acceptability among domestic helpers. Interestingly, we found they have strong preference for onsite training over online participation. This suggests that face-to-face engagement remains highly valued, even among populations with access to digital tools. While online delivery offers flexibility, these results highlight the need for a hybrid approach that combines digital resources with in-person sessions to maximize reach and effectiveness. This preference may reflect the social and motivational benefits of group-based activities, which are difficult to replicate in virtual formats.

The observed behavioral changes were concentrated in greater intake of healthy foods, such as fruits, vegetables, and breakfast, rather than reductions in snack consumption. This pattern suggests that interventions emphasizing positive additions to diet may be more culturally acceptable than those focused on restriction.

The finding that over 95% of participants expressed willingness to recommend the program to the families they serve points to an important opportunity for community-level health promotion. Domestic helpers often influence household food choices and caregiving practices; thus, empowering them with health knowledge may have spillover effects that extend beyond individual participants to the broader family environment. Leveraging this dynamic could amplify the public health impact of such interventions.

This study has several strengths. First, the intervention was designed to be pragmatic and context-specific, incorporating hybrid delivery and standing-posture yoga to accommodate the unique work conditions of domestic helpers. Second, our study has relatively large sample size, allowing us to conduct stratified analyses by baseline health behaviors and mental stress. Third, in addition to questionnaire surveys, we also collected qualitative feedback. The latter is consistent with the findings from surveys, showing high feasibility and acceptability of the intervention.

However, several limitations should be acknowledged. The pre-post design without a control group limits causal inference and introduces potential biases such as confounding and regression to the mean.^24^ Outcomes were assessed using self-reported measures, which are subject to recall and social desirability bias.^25^ Our intervention period is relatively short. Future studies using randomized designs, objective measures of diet and activity, and longer intervention, would be worthwhile.

Our findings have important implications for practice and policy. Domestic helpers occupy a pivotal yet precarious position in household care ecosystems. A targeted, low-cost, low-burden intervention that demonstrably improves health behaviors and mental well-being in the short term is actionable for employers, agencies, and public health regulators. The hybrid model and standing-posture yoga are particularly suited to live-in arrangements, suggesting scalability across districts and potentially across the broader in Asia-Pacific region. Integrating such programs into Sunday community hubs, NGO partnerships, or employer-supported wellness initiatives may enhance reach and sustainability. Given the stronger effects among those with worse baseline status, screening-guided targeting could optimize impact. Policymakers might also consider protected time for health promotion on rest days and micro-grants to community groups delivering such interventions.

## Data Availability

All data produced in the present study are available upon reasonable request to the authors after publication.

## Funding

This project was supported by Knowledge Exchange Funding (KE-IP-2024/25-66), funded by the University of Hong Kong.

## Acknowledgement

We gratefully acknowledge the funding support from the Knowledge Exchange Funding of the University of Hong Kong. We extend our special thanks to Prof. Ching-Lung Cheung for helping with the funding application, and Dr. Tiffany Leung for helping with website maintenance and the preparation of educational material. We also thank Ms. Yuanqing Xia, Ms. Xin Huang, Dr. Wenming Shi, and Ms Junmeng Zhang for assistance with fieldwork. We are appreciative of the employment agencies (WEcarehelper and Fair Employment Agency Limited) and the NGO (HELP for Domestic Helpers) that supported participant recruitment. Our sincere thanks go to all participants for joining this project and providing valuable feedback. Finally, we thank *Ming Pao* for reporting our project.

## Notes

### Competing Interest Statement

The authors have declared no competing interest.

### Author Declarations

This study was approved by Institution Review Board of The University of Hong Kong/ Hospital Authority Hong Kong West Cluster (approval number UW 24-823).

